# Predictors of Intense Treatment in the Emergency Department Among Older Adults With Serious Life-Limiting Illnesses: A Five-Year Cross-sectional Analysis of Medicare Claims Data

**DOI:** 10.1101/2023.06.19.23291571

**Authors:** Oluwaseun Adeyemi, Nina Siman, Keith Goldfeld, Jacob Hill, Allison Cuthel, Charles DiMaggio, Joshua Chodosh, Corita Grudzen

## Abstract

**Background:** Treatment intensity of end-of-life care is the degree of aggressiveness of medical care aimed at providing life-prolonging medical care to patients with serious life-limiting illnesses. This study aims to assess the demographic and health characteristics associated with older adults with serious life-limiting illnesses who received highly intense end-of-life care.

**Methods:** For this cross-sectional analysis, we pooled the 2015 to 2019 Medicare claims data of adults 65 years and older who visited at least one of the 29 emergency departments (EDs) enrolled in the Primary Palliative Care for Emergency Medicine. We identified those with serious life-limiting illnesses using a Gagne score of seven or higher. Our outcome measure was treatment intensity, defined using acute care and intensive care unit (ICU) admissions. Acute care admission was measured as a binary variable and ICU admission was measured as a three-point nominal variable. The predictor variables were age, sex, race/ethnicity, and illness severity (Gagne score). To assess the odds of acute care and ICU admissions, we used a generalized estimating equation model and a multinomial regression model, respectively. We performed the same analyses among the population without serious life-limiting illnesses to observe differences in effect sizes of intense treatment.

**Results:** Of the 301,083 older adults that visited one of the 29 EDs, 13% had serious life-limiting illnesses. Age was associated with 9% and 7% increased odds of acute care (95% CI: 1.04 – 1.14) and ICU (95% CI: 1.02 – 1.12) admissions. We reported significant associations by sex, race/ethnicity, and illness severity (Gagne score). The effect sizes of the observed association between measures of treatment intensity and the demographic and health characteristics were smaller among those with serious life-limiting illnesses compared to those without serious life-limiting illnesses.

**Conclusion:** Older adults with serious life-limiting illnesses who present to the ED experience intense treatment. Identifying demographic and health characteristics associated with treatment intensity may inform the need for serious illness conversations in the ED.

## Introduction

Within the last month of life, half of United States (U.S.) older adults with serious life-limiting illnesses visit the Emergency Department (ED).^1^ These serious life-limiting illnesses mostly include advanced cancer, heart failure, chronic obstructive pulmonary disease (COPD), and end-stage kidney disease.^2,3^ ED visits represent inflection points in the chronic illness trajectory of patients with serious life-limiting illnesses and the care they receive in the ED may impact the quality of their end-of-life.^4,5^ The ED is typically geared towards providing highly intense, life-sustaining care,^6^ and such high-intensity treatment may be inconsistent with the desired quality of end-of-life care of patients with serious life-limiting illnesses.^7^

Treatment intensity of end-of-life care is defined as the degree of aggressiveness of medical care and the rate of utilization of healthcare resources aimed at providing life-prolonging medical care to dying patients within a specified time frame before demise.^8,9^ Measures of highly intense end-of-life care typically include rates of acute care and intensive care unit (ICU) admissions, length of in-hospital and ICU stays, exposure to invasive procedures, placement on life support, and in-hospital death.^8,9^ Less intense end-of-life care include measures of comfort care and palliative care.^10-12^ Earlier studies have reported that highly intensive therapies do not result in quality end-of-life care.^13-15^ Additionally, patients with serious life-limiting illnesses tend to prefer having their care delivered at home and dying at home.^16,17^

Identifying the patient characteristics associated with exposure to highly intense end-of-life treatment may provide information on the population at risk of reduced quality of end-of-life care. Additionally, understanding the demographic and health characteristics of patients who receive highly intense end-of-life care may aid in the identification of the patient population that will benefit the most from educational interventions aimed at providing knowledge on making informed decisions on end-of-life care. No study has assessed the level of intense treatment patients with life-limiting illnesses who visit the ED receive. We, therefore, aim to assess the demographic and health characteristics associated with older adults (65 years and older) with serious life-limiting illnesses who received highly intense end-of-life care following an index ED visit and assess how the strength of the association varies among older adults without serious life-limiting illnesses.

## Methods

### Study Design and Population

For this cross-sectional analysis, we first examined the pattern of healthcare utilization among older adults who visit the ED. Thereafter, we stratified the population into those with and without serious life-limiting illnesses to gain an understanding of how the pattern of healthcare utilization differed across the group and how intensely older adults with serious life-limiting illnesses are managed. Our study population was older adults who visited one of the 29 EDs involved in a large national study entitled, “Primary Palliative Care for Emergency Medicine,” (PRIM-ER). The PRIM-ER study is a pragmatic cluster-randomized stepped-wedge clinical trial aimed at assessing how primary palliative care delivery in the ED affects healthcare utilization among patients with serious life-limiting illnesses. Additional information on the study’s protocol has been published, ^18-21^ and the study is registered on Clinical Trials.gov (NCT03424109).^22^ For this index study, our focus was to examine the intensity of care before the start of the PRIM-ER interventions across 29 EDs.

### Data Source

Our study used Medicare claim records accessed through the Centers for Medicare and Medicaid Services Chronic Conditions Data Warehouse. The Medicare claims data is the largest US healthcare administrative database that provides health information on individuals 65 years and older and those with disabilities who are less than 65 years.^23^ We examined the inpatient, ambulatory, and carrier claim files as well as the beneficiary summary file ABCD. We merged these files using unique beneficiary identifiers.

### Inclusion and Exclusion Criteria

Since the PRIM-ER intervention started in April 2019, we pooled Medicare claims data between April 1, 2015 - March 31, 2019. Between the selected period, there were a total of 301,047 unique Medicare enrollees, aged 65 years and older, who had an index visit at one of the 29 EDs. We identified older adults with serious life-limiting illnesses by computing the Gagne index, a measure of illness severity and six-month mortality (discussed below).^24^ Older adults with a Gagne index higher than six were classified as having serious life-limiting illnesses. Older adults with a Gagne index of six or less were classified as not having serious life-limiting illnesses. Our final analytic sample, therefore, consisted of older adults with serious life-limiting illnesses (n=38,793) and those without serious life-limiting illnesses (n=262,251).

### Outcome Measures

Our main outcome variables were two measures of treatment intensity: acute care and ICU admission. We selected these variables consistent with earlier literature that defined how treatment intensity among patients with life-limiting illness are measured.^8,9^ Acute care admission, defined as any ED disposition that led to inpatient admission, was measured as a binary variable. Also, ICU admission, defined as any ED disposition that led to admission into the ICU unit, was measured as a three-point nominal variable – admitted to ICU, admitted but not to ICU, and not admitted.

### Predictor Variables

The predictor variables were demographic and health characteristics. The demographic characteristics include age, sex, and race/ethnicity. Age was measured as a continuous variable, sex was measured as a binary variable, and race/ethnicity was measured as a four-level categorical variable – non-Hispanic White, non-Hispanic Black, Hispanic, and other races. The health characteristic we selected was the Gagne index.^24^ The Gagne index was adapted from the Romano-Charlson Index and the Elixhauser system and was developed to predict one-year mortality in community-dwelling older adults.^24^ The index is calculated based on the presence or absence of 20 diagnoses (Appendix 1), which we identified using their ICD-9 (January 2014-September 2015) and ICD-10 (October 2015-December 2018) codes from the inpatient and ambulatory claims. We measured the Gagne index as a continuous variable, with the scores ranging from -3 to 25. Since we used the Gagne index as the stratification variable, the Gagne index value for those with serious life-limiting illnesses is 7 to 25. Lastly, our study population was clustered across the 29 EDs. We, therefore, used the EDs as a random effect measure to control for institutional-level characteristics.

### Human Subject Concern

This study was approved by the NYU Grossman School of Medicine Institutional Review Board (Study ID i18-00607).

### Statistical Analyses

We used descriptive statistics to characterize demographic and health variables. To estimate the association between the predictors and acute care admission, we used a generalized estimating equation (GEE) model. Specifically, we used a one-step GEE algorithm that can accommodate the extremely large cluster sizes in our study.^25^ First, we ran the model on the entire study population and thereafter, we ran the model separately for those with and without serious life-limiting illnesses. We reported the adjusted odds (and 95% confidence interval (95% CI)) of acute care admission across the entire study population and separately among those with and without serious life-limiting illnesses. Also, to estimate the association between the predictors and ICU admission, we performed a multinomial regression analysis. We reported the adjusted odds (and 95% CI) of ICU admission across the population with serious life-limiting illnesses. Analyses were conducted using SAS Version 7.1 ^26^ and R/Databricks ^27,28^, both accessed in the Chronic Conditions Warehouse Virtual Research Data Center.

## Results

Table 1 includes the characteristics of our study sample (N=301,083) and those with (n=38,793; 12.9%) and without (n=262,251; 87.1%) serious life-limiting illnesses. The mean (SD) age across the study population was 76.1 (8.1) years and the majority were females (54%), and non-Hispanic White (77.4%). The mean (SD) Gagne score was 2.4 (3.3). Across the entire study population, 46% had acute care admission and 6% had ICU admission. The population that experienced acute care was older (mean age: 77.1 vs 75.2), with a higher mean Gagne index (3.1 vs. 1.9) (Table 2). Also, the Gagne index increased stepwise from those who were not admitted (1.9), to those admitted but not to ICU (3.0), and those admitted to ICU (3.3).

**Table 1:** Summary of the demographic and health characteristics of the study population from the Medicare Claims data stratified into those with and without serious life-limiting illnesses.

| Variables | All Patients<br>(N=301,044) | Without Serious Life-limiting Illness<br>(n=262,251 (87.1%)) | With Serious Life-Limiting Illness<br>(n=38,793 (12.9%)) |
| --- | --- | --- | --- |
| Age (Mean, SD) | 76.1 (8.1) | 76.0 (8.1) | 76.9 (8.1) |
| Sex (N, %) |  |  |  |
| Female | 163,138 (54.2) | 144,639 (55.2) | 184,499 (47.7) |
| Male | 137,906 (45.8) | 117,612 (44.8) | 20,294 (52.3) |
| Race/Ethnicity (N, %) |  |  |  |
| Non-Hispanic White | 233,018 (77.4) | 202,914 (77.4) | 30,104 (77.6) |
| Non-Hispanic Black | 39,734 (13.2) | 33,999 (13.0) | 5,735 (14.8) |
| Hispanic | 5,118 (1.7) | 4,611 (1.8) | 507 (1.3) |
| Other Race* | 23,174 (7.7) | 20,727 (7.9) | 2,447 (6.3) |
| Gagne Score (Mean, SD)** | 2.4 (3.3) | 1.4 (1.9) | 9.3 (2.3) |
| Acute Care (N, %) |  |  |  |
| Yes | 138,540 (46.0) | 113,780 (43.4) | 24,760 (63.8) |
| No | 162,504 (54.0) | 148,471 (56.6) | 14,033 (36.2) |
| ICU Admission (N, %) |  |  |  |
| Admitted to ICU | 17,402 (5.8) | 13,788 (5.3) | 3,614 (9.3) |
| Admitted, not ICU | 121,138 (40.2) | 99,992 (38.1) | 21,146 (54.5) |
| Not Admitted | 162,504 (54.0) | 148,471 (56.6) | 14,033 (36.2) |

Patients with serious life-limiting illnesses are defined as having a Gagne score higher than six
\*Other races include Asian, North American Native, Other, and Unknown
\*\*Gagne Score: A combined comorbidity score computed from 20 diagnoses.
ICU: Intensive Care Unit

**Table 2:** Frequency distribution and summary statistics of the demographic and health characteristics stratified by acute care admission or Intensive Care Unit (ICU) admission status.

| Variables | Acute Care Admission |  | ICU Admission |  |  |
| --- | --- | --- | --- | --- | --- |
|  | Yes<br>(n=138,540; 46.0%) | No<br>(n=162,504; 54.0%) | Admitted to ICU<br>(n=17,403; 5.8%) | Admitted, Not ICU<br>(n=121,138; 40.2%) | Not Admitted<br>(n=162,504; 54.0%) |
| Age (Mean, SD) | 77.1 (8.4) | 75.2 (7.7) | 76.6 (8.1) | 77.2 (8.4) | 75.2 (7.7) |
| Sex (N, %) |  |  |  |  |  |
| Female | 70,660 (51.0) | 92,478 (56.9) | 7,872 (45.2) | 62,788 (51.8) | 92,478 (56.9) |
| Male | 67,880 (49.0) | 70,026 (43.1) | 9,530 (54.8) | 58,350 (48.2) | 70,026 (43.1) |
| Race/Ethnicity (N, %) |  |  |  |  |  |
| Non-Hispanic White | 110,433 (79.7) | 122,585 (75.4) | 13,664 (78.5) | 96,769 (79.9) | 122,585 (75.4) |
| Non-Hispanic Black | 16,159 (11.7) | 23,575 (14.5) | 1,941 (11.2) | 14,218 (11.7) | 23,575 (14.5) |
| Hispanic | 1,958 (1.4) | 3,160 (1.9) | 211 (1.2) | 1,747 (1.4) | 3,160 (1.9) |
| Other Race* | 9,990 (7.2) | 13,184 (8.1) | 1,586 (9.1) | 8,404 (6.9) | 13,184 (8.1) |
| Gagne Score (Mean (SD)) | 3.1 (3.6) | 1.9 (2.9) | 3.3 (3.9) | 3.0 (3.6) | 1.9 (2.9) |

\*Other race includes Asian, North American Native, Other, and Unknown

There were differences in the demographic and health characteristics among those with and without serious life-limiting illnesses (Table 1). The population with serious life-limiting illnesses was slightly older than those without serious life-limiting illnesses (76.9 years vs. 76.0 years). Also, those with serious life-limiting illnesses were predominantly males (52%) while those without serious life-limiting illnesses were predominantly females (55%). The proportion of Blacks with and without serious life-limiting illnesses was 15% and 13%, respectively. Also, 64% and 9% of those with serious life-limiting illnesses had acute care and ICU admission, respectively, compared to 43% and 5% of those without serious life-limiting illnesses.

Table 3 shows the results of the GEE model. A unit increase in age was associated with 29% increased odds of acute care admission (AOR: 1.29; 95% CI: 1.24 – 1.33). Males were 27% more likely to be admitted to acute care compared to females (AOR: 1.26, 95% CI 1.25 – 1.29). Also, non-Hispanic Blacks (AOR 0.84, 95% CI 0.78 – 0.90), Hispanics (AOR 0.69, 95% CI 0.60 – 0.78), and those other races (AOR 0.93, 95% CI 0.88 – 0.99) were less likely to be admitted to acute care compared to non-Hispanic Whites. A unit increase in the Gagne index was associated with 11% increased odds of acute care admission (AOR: 1.11; 95% CI: 1.09 – 1.12).

**Table 3:** Association between the demographic and health characteristics and acute care admission among those with and without serious life-limiting illnesses.

| Variables | Acute Care Admission (Adjusted OR (95% CI)) |  |  |
| --- | --- | --- | --- |
|  | All Patients | Without Serious Life-limiting Illness | With Serious Life-limiting Illness |
| Age | **1.29 (1.24, 1.33)** | **1.30 (1.26, 1.35)** | **1.09 (1.04, 1.14)** |
| Sex |  |  |  |
| Female | Ref | Ref | Ref |
| Male | **1.26 (1.25, 1.29)** | **1.30 (1.28, 1.32)** | **1.07 (1.03, 1.11)** |
| Race/Ethnicity |  |  |  |
| Non-Hispanic White | Ref | Ref | Ref |
| Non-Hispanic Black | **0.84 (0.78, 0.90)** | **0.83 (0.77, 0.89)** | **0.86 (0.78, 0.93)** |
| Hispanic | **0.69 (0.60, 0.78)** | **0.66 (0.58, 0.74)** | 0.94 (0.76, 1.15) |
| Other Race | **0.93 (0.88, 0.99)** | **0.93 (0.87, 0.99)** | 0.95 (0.87, 1.03) |
| Gagne Score | **1.11 (1.09, 1.12)** | **1.11 (1.10, 1.13)** | **1.06 (1.04, 1.09)** |

Significant association in bold.
Patients without serious life-limiting illnesses are defined as having a Gagne score of six or less.
Patients with serious life-limiting illnesses are defined as having a Gagne score higher than six.

Among those with serious life-limiting illnesses, a unit increase in age was associated with a 9% increased odds of acute care admission (AOR: 1.09; 95% CI: 1.04 – 1.14) (Table 3). Males were 7% more likely to be admitted to acute care compared to females (AOR: 1.07, 95% CI 1.03 – 1.11). Also, non-Hispanic Blacks were 14% less likely to be admitted to acute care compared to non-Hispanic Whites (AOR 0.86, 95% CI 0.78 – 0.93). A unit increase in the Gagne index was associated with a 6% increased odds of acute care admission (AOR: 1.06; 95% CI: 1.04 – 1.09). Similarly, a unit increase in age was associated with a 7% increased odds of ICU admission (AOR: 1.07; 95% CI: 1.02 – 1.12) (Table 4). Males were 20% more likely to have ICU admission compared to females (AOR: 1.20, 95% CI 1.12 – 1.30). Also, non-Hispanic Blacks were 21% less likely to be admitted to acute care compared to non-Hispanic Whites (AOR 0.79, 95% CI 0.71 – 0.87). A unit increase in the Gagne index was associated with an 11% increased odds of acute care admission (AOR: 1.11; 95% CI: 1.09 – 1.13).

**Table 4:** Association between the demographic and health characteristics and intensive care unit (ICU) admission among patients with serious life-limiting illnesses.

| Variables | ICU Admission (Adjusted OR (95% CI)) |  |
| --- | --- | --- |
|  | Admitted to ICU | Admitted, Not ICU |
| Age | **1.07(1.02, 1.12)** | **1.14(1.11, 1.17)** |
| Sex |  |  |
| Female | Ref | Ref |
| Male | **1.20 (1.12, 1.30)** | **1.04 (1.00, 1.09)** |
| Race/Ethnicity |  |  |
| Non-Hispanic White | Ref | Ref |
| Non-Hispanic Black | **0.79(0.71, 0.87)** | **0.75(0.91, 0.80)** |
| Hispanic | 0.93(0.67, 1.29) | 0.95(0.78, 1.14) |
| Other | 1.04(0.90, 1.20) | 0.92(0.84, 1.00) |
| Gagne Score | **1.11(1.09, 1.13)** | **1.06(1.05, 1.07)** |

Significant association in bold.

## Discussion

In this study, we defined treatment intensity for patients with serious life-limiting illnesses using acute care and ICU admissions. These two measures are the most widely used metrics of treatment intensity.^8,29^ Our first aim was to assess the factors associated with treatment intensity among older adults with serious life-limiting illnesses. Our result showed that increasing age and worsening illness severity are associated with intense treatment among older patients that present in the ED. Also, males and non-Hispanic Whites with serious life-limiting illnesses are more likely to experience more intense treatment. Our second aim was to compare how the effect sizes of the factors associated with treatment intensity differed among patients with and without serious life-limiting illnesses. Our result showed that while age, sex, race/ethnicity, and Gagne index exhibited similar patterns of association among the population with and without serious life-limiting illnesses, the effect sizes were smaller among the patient population with serious life-limiting illnesses. While the smaller effect size of treatment intensity in the patient population with serious life-limiting illnesses suggests a step down in the aggressiveness of treatment, the strong association between age, sex, race/ethnicity, and Gagne index suggest that there is room for improvement.

Our results showed that age was associated with increased odds of intense treatment among older adults with serious life-limiting illnesses. Considering that the population classified as having serious life-limiting illnesses had an estimated six months or less to live, it is debatable if such ICU admission these older adults experienced were of any benefit. While physician characteristics and local practice patterns may influence the decision to provide intense care to patients with serious life-limiting illnesses,^30-34^ patients and family determinants equally exist. Earlier studies have reported that physicians’ race/ethnicity, years of practice, and attitudes toward end-of-life care influence the decision to provide life-prolonging treatments for dying patients. ^31-34^ Also, patients and family or caregivers may request life-prolonging treatment despite a lack of evidence of its benefit.^30^ Our study further shows that the more severe the illness patients with serious life-limiting illnesses have, the more the likelihood to engage such a patient to have intense treatment. The severity of an illness for a patient with serious life-limiting illnesses should present the opportunity for the providers to initiate serious illness conversations. However, if serious illness conversations are only introduced only when patients with life-limiting illnesses present with severe illness, such conversations might be misconstrued as the emergency provider’s unwillingness to save lives. Engaging in serious illness conversations and encouraging patients with serious life-limiting illnesses to have a documented preference for care early in the illness trajectory may benefit the patient, provider, and the healthcare system.

In our study, serious life-limiting illnesses were disproportionately higher among male older adults compared to females. Additionally, among the population with serious life-limiting illnesses, males were likely to experience intense treatment compared to females. This observed sex difference in treatment intensity supports findings from prior studies.^35-37^ Earlier studies have reported that females with serious life-limiting illnesses were less likely to prefer life-prolonging or aggressive treatment and are more likely to have a do-not-resuscitate order.^35-37^ Yet, it appears that sex differences in the receipt of intense end-of-life treatment are amenable to change. Sharma and colleagues reported that although men with metastatic cancer were three times more likely to receive ICU care compared to females, men who reported having end-of-life discussions were 74 percent less likely to receive unnecessary ICU care compared to men who did not report having end-of-life discussions.^38^ Engaging males in serious illness conversations is however not a linear process. Male older adults with serious life-limiting illnesses were more likely to avoid discussion around death and are less likely to recognize that their illnesses are incurable.^39-41^ Initiating serious illness conversations, especially for men with serious life-limiting illnesses, may be more successful if such conversations are personalized, the providers have a prior understanding of the sociocultural motivation for life-prolonging treatment, and there exists some level of trust between the patient and provider. Additionally, serious illness conversations should be excellently timed to capture those periods when patients may be more receptive to the hard facts about their illness trajectory, and providers must understand to what extent such patients will permit family and friends to share in the end-of-life decision-making process.

Earlier studies have reported that compared to Blacks and Hispanics were more likely to opt for life-prolonging treatment and less likely to have end-of-life care instructions compared to Whites.^42-45^ Contrastingly, a few other studies have reported that Whites with serious life-limiting illnesses were more likely to have more acute care and ICU admissions compared to Blacks, especially in their terminal admissions. ^46,47^ Although our study showed that Blacks were less likely to receive intense end-of-life care, racial and ethnic differences in end-of-life care should be contextualized rather than generalized. While regional differences in end-of-life treatment intensity exist, sociocultural and religious beliefs may explain these observed differences and may account for the conflicting results. In the most ideal settings, there should not be racial or ethnic differences in end-of-life care and centers and regions where those differences exist, a deeper understanding of the root cause of such disparities should be explored and managed. We opine that all patients with serious life-limiting illnesses should be engaged in serious illness conversations and the treatment they receive should be consistent with their goals of care.

This study has its limitations. It is a cross-sectional study and we cannot assume causality. Our measures of treatment intensity are limited to billable processes. Hence, we cannot capture evidence of serious illness conversations, which is a potential confounder of the associations we evaluated. Also, we are unable to determine whether the care received was consistent with patient goals since such information is unavailable in the Medicare claims data. Other sociodemographic characteristics such as education, marital status, income, presence of caregivers, family support, patient-reported treatment outcomes, and quality of life are unavailable in the Medicare claims data. These characteristics may influence the strength of the association we report. Additionally, our study is limited to patients who visited at least one of the 29 EDs involved in the PRIM-ER study. Hence, the result of our findings has limited generalizability. Despite these limitations, our study is strengthened by its large sample size and the use of Medicare claims data, which is a valuable resource for epidemiologic and health services research.^26^ It is, therefore, possible that each institution can apply our methodology to assess how intensely they manage patients with serious life-limiting illnesses and which unique factors apply to their settings. Additionally, this study creates the platform to assess the efficacy of pragmatic interventions aimed at implementing palliative care services in the ED. By observing how the treatment intensity for patients with serious life-limiting illnesses differed before and during the years of intervention, we will be able to contribute to policy guidelines aimed at improving access to palliative care services in the ED.

## Conclusion

Older adults with serious life-limiting illnesses who present to the ED experience intense treatment. The likelihood of intense treatment increases with increasing age and illness severity. Additionally, males and non-Hispanic Whites are at higher odds of experiencing intense end-of-life treatment. Interventions aimed at improving serious illness conversations in the ED may reduce the likelihood of treatments that may not be of benefit to patients with serious life-limiting illnesses.

## Data Availability

All data produced in the present study are available upon reasonable request to the authors

